# Impact of Left Ventricular Venting on Acute Brain Injury in Patients with Cardiogenic Shock: An Extracorporeal Life Support Organization Registry Analysis

**DOI:** 10.1101/2024.11.20.24317676

**Authors:** Shi Nan Feng, Winnie L. Liu, Jin Kook Kang, Andrew Kalra, Jiah Kim, Akram Zaqooq, Melissa A. Vogelsong, Bo Soo Kim, Daniel Brodie, Patricia Brown, Glenn J. R. Whitman, Steven Keller, Sung-Min Cho

**Author notes:** **Corresponding Author** Sung-Min Cho, DO, MHS, Divisions of Neurosciences Critical Care and Cardiac Surgery The Johns Hopkins Hospital, 600 N. Wolfe Street, Phipps 455, Baltimore, MD 21287. S.N.F. and W.L.W. contributed equally as co-first authors.

## Abstract

**Background:** While left ventricular (LV) venting reduces LV distension in cardiogenic shock patients on venoarterial extracorporeal membrane oxygenation (VA-ECMO), it may also amplify risk of acute brain injury (ABI). We investigated the hypothesis that LV venting is associated with increased risk of ABI. We also compared ABI risk of the two most common LV venting strategies, percutaneous microaxial flow pump (mAFP) and intra-aortic balloon pump (IABP).

**Methods:** The Extracorporeal Life Support Organization registry was queried for patients on peripheral VA-ECMO for cardiogenic shock (2013-2024). ABI was defined as hypoxic-ischemic brain injury, ischemic stroke, or intracranial hemorrhage. Secondary outcome was hospital mortality. We compared no LV venting with 1) LV venting, 2) mAFP, and 3) IABP using multivariable logistic regression. To compare ABI risk of mAFP vs. IABP, propensity score matching was performed.

**Results:** Of 13,276 patients (median age=58.2, 69.9% male), 1,456 (11.0%) received LV venting (65.5% mAFP and 29.9% IABP), and 525 (4.0%) had ABI. After multivariable regression, LV-vented patients had increased odds of ABI (adjusted odds ratio (aOR)=1.76, 95% CI=1.29, 2.37, p<0.001) but no difference in mortality (aOR=1.08, 95% CI=0.91-1.28, p=0.39) compared to non-LV-vented patients. In the propensity-matched cohort of IABP (n=231) vs. mAFP (n=231) patients, there was no significant difference in odds of ABI (aOR=1.35, 95%CI=0.69-2.71, p=0.39) or mortality (aOR=0.88, 95%CI=0.58-1.31, p=0.52).

**Conclusions:** LV venting was associated with increased odds of ABI but not mortality in patients receiving peripheral VA-ECMO for cardiogenic shock. There was no difference in odds of ABI or mortality for IABP vs. mAFP patients.

**Clinical Perspective:** In patients receiving peripheral venoarterial extracorporeal membrane oxygenation (VA-ECMO) for cardiogenic shock, left ventricular venting is associated with increased odds of acute brain injury (ABI) but not mortality. However, mode of venting—intra-aortic balloon pump (IABP) or percutaneous microaxial flow pump (mAFP)—does not appear to impact either odds of ABI or mortality. These findings highlight a link between venting strategies and neurological outcomes in this high-risk population. Clinicians must weigh the benefits of venting against ABI risk when managing neurocritically ill patients, though our findings provide reassurance clinicians that both IABP and mAFP may offer comparable neurologic safety profiles.

## INTRODUCTION

Cardiogenic shock is a life-threatening condition characterized by low cardiac output, end-organ hypoperfusion, and high mortality.^1,2^ In recent years, venoarterial extracorporeal membrane oxygenation (VA-ECMO) has been increasingly employed as a short-term rescue strategy in patients with cardiogenic shock, offering partial or full hemodynamic and respiratory support while reducing myocardial workload.^3^ However, this has not always translated to reduced mortality for patients.^4^

Notably, the ECMO circuit can impose additional strain on the left ventricle (LV) by increasing afterload and altering normal blood flow, and left ventricular (LV) distension is a serious complication that can occur in up to 60% of patients.^5–8^ This weakened ejection can lead to blood pooling, elevated LV pressures, and increased risk of pulmonary edema, pulmonary hemorrhage, myocardial ischemia, cerebral hypoxia, and LV failure.^9^ Given these risks, dual mechanical support using a secondary device for LV venting has been explored to enhance both systemic circulation and LV function.^10^ LV venting is a critical intervention in patients receiving VA-ECMO that works by offloading intraventricular pressure from the LV.^7,9,11^ Currently, the two most common mechanical LV venting devices are the percutaneous microaxial flow pump (mAFP) and the intra-aortic balloon pump (IABP). The mAFP is a catheter-based microaxial rotary pump that actively pumps blood from the LV to the ascending aorta, lowering LV pressure and myocardial wall stress.^12^ The IABP inflates during diastole to augment coronary blood flow and deflates before systole, reducing afterload and assisting the LV in ejecting blood.^5,13,14^

While LV venting can mitigate the risk of myocardial damage, it can also introduce additional risks. Studies suggest that LV venting may increase risk of acute brain injury (ABI), which can occur in up to 11-20% of VA-ECMO patients and represents a leading cause of mortality.^15^ In particular, studies have associated the use of mAFP for LV unloading during VA-ECMO (ECMELLA) with higher rates of complications including ABI compared to VA-ECMO alone, though findings are mixed regarding whether ischemic stroke or ICH risk is elevated.^13,16^ Additionally, studies have associated lower pulse pressures with increased risk of ABI in VA-ECMO patients, which may impact ABI risk of different LV venting strategies.^17^ Despite these risks and the increasing use of LV venting in clinical practice, the complex interplay between circulatory support devices, cerebral perfusion, and the risk of ABI remains poorly understood. Moreover, there is a lack of clarity on which VA-ECMO patients should receive LV venting despite the risk of complications.^14^

This study aims to characterize the association between LV venting and ABI in patients with cardiogenic shock receiving peripheral VA-ECMO. In comparing the effects of mAFP and IABP on ABI outcomes using the largest registry of ECMO patients globally, our work seeks to clarify the impact of LV venting strategies on ABI risk and improve clinical outcomes for this complex population.

## METHODS

### Patients

We conducted a retrospective analysis of the ELSO registry for patients who received peripheral VA-ECMO for cardiogenic shock from January 1, 2013 – June 21, 2024. We excluded patients treated with VV-ECMO, patients who were centrally cannulated, pediatric patients (<18), patients who received more than one ECMO run, patients who had conversions in ECMO mode, and patients with missing demographic, LV venting, or ABI data.

Patients were sub-grouped by LV venting or no LV venting. LV venting was defined using Current Procedural Terminology (CPT) codes^18^ **(SUPPLEMENTAL TABLE I)**. Procedure timing for LV-vented patients was limited to “On-ECLS”. LV venting procedure timing that was “Pre-ECLS” but within 1 hour of ECMO cannulation was also included as “On-ECLS”. Otherwise, patients who received LV venting “Pre- ECLS” or “Post-ECLS” were categorized as no LV venting since the procedure did not occur during VA-ECMO treatment.

This retrospective observational cohort study was approved by the Johns Hopkins Hospital Institutional Review Board with a waiver of informed consent (IRB00216321) and reported using Strengthening the Reporting of Observational Studies in Epidemiology (STROBE) guidelines.^19^

### Data Source

The Extracorporeal Life Support Organization (ELSO) registry is a voluntary international database that collects information on use, indications, complications, and outcomes of ECMO support in adults and children from more than 50 countries.^20^ Diagnosis and medical history are reported according to the International Classification of Diseases 9^th^ edition (ICD-9) and 10^th^ edition (ICD-10) codes.

For all included patients, we extracted the following information from the ELSO registry database: pre-ECMO demographic information; pre-ECMO clinical variables; laboratory values; on-ECMO clinical variables including LV venting procedures; and ECMO-associated morbidity and mortality, including renal replacement therapy (RRT), hemolysis, arrythmia, gastrointestinal hemorrhage, and ABI.

### Definitions

ABI was defined as hypoxic-ischemic brain injury (HIBI), ischemic stroke (IS), and intracranial hemorrhage (ICH) including intraventricular hemorrhage. In the ELSO registry, IS is defined as central nervous system (CNS) infarction determined by ultrasound, computed tomography (CT), or magnetic resonance imaging (MRI). ICH is defined as intra- or extra-parenchymal CNS hemorrhage or intraventricular CNS hemorrhage determined by ultrasound, CT, or MRI. HIBI is defined as CNS diffuse ischemia.

LV venting strategies were grouped into 3 categories: mAFP, IABP, and Other LV Venting. MAFP included all Impella CPT codes **(SUPPLEMENTAL TABLE I)**. Other LV Venting included closed heart atrial septostomy, open heart atrial septostomy with cardiopulmonary bypass, insertion of left heart vent by thoracic incision, insertion of catheter into right pulmonary artery, and transvenous atrial septectomy or septostomy with balloon including cardiac catheterization. Cardiogenic shock was defined as ICD-9 code 785.51 and ICD-10 code R57.0.

Arterial blood gases were collected at baseline/pre-ECMO and at 24 hours, and PaCO_2_ difference was defined as PaCO_2_ at 24 hours - PaCO_2_ at baseline/pre-ECMO. The pre-ECLS hemodynamics and arterial blood gas (ABG) values were measured no more than 6 hours before ECLS. Twenty-four– hour ABG values were drawn between 18 and 30 hours after ECLS start time. RRT occurred during ECMO support.

### Outcomes

The primary outcome was ABI during ECMO support. ABI outcome was assigned if the injury occurred during ECMO support and after the LV venting procedure time. The secondary outcome was in-hospital mortality.

### Statistical Analysis

Patient characteristics, clinical management, and outcomes data were summarized as medians and interquartile range [IQR] for continuous variables. Numbers and percentages were calculated for categorical variables. Continuous baseline characteristics were compared using the Wilcoxon rank-sum test, and discrete characteristics were compared using the Chi-square test. Normality of variables was assessed using Shapiro-Wilk testing and histogram visualization. A p-value of <0.05 was considered to be statistically significant. All statistical analyses were performed using R Studio (R 4.1.2, 2022).

The association between LV venting and ABI was examined using multivariable logistic regression to balance for clinically pre-selected risk factors. The use of 1) LV venting vs. no LV venting, 2) mAFP vs. no LV venting, and 3) IABP vs. no LV venting was compared. To examine the risk of ABI for patients receiving mAFP vs. IABP, propensity-score matching was performed using 1:1 nearest neighbor matching without replacement within a caliper width of 0.2, with IABP as the as the dependent variable. Listwise deletion of cases with missing covariates or independent variables was used. Satisfactory matching was defined as an absolute value of the standardized mean difference (SMD) of <0.10. Propensity scores were obtained by logistic regression. Participants were matched by variables recorded in the ELSO registry including age, sex, BMI, hours on ECMO, pre-ECMO pH, pre-ECMO PaO_2_, PaO_2_ at 24 hours, PaCO_2_ difference, lactate at 24 hours post-ECMO cannulation, pre-ECLS cardiac arrest, pump flow at 4 hours post-ECMO cannulation, on-ECMO RRT, and on-ECMO complications including gastrointestinal hemorrhage, arrythmia, and hemolysis. Covariate selection for multivariable models was guided by both literature review and clinical relevance of candidate predictors.

After matching, multivariable logistic regression was used to compare ABI risk for mAFP vs. IABP groups. In our analyses comparing mAFP vs. IABP, mAFP support was used as the reference group given that it was the most frequently used type of LV venting in our study population. Odds ratios (ORs) with 95% confidence intervals (CIs) were calculated. Collinearity between confounders was assessed, with a variance inflation factor (VIF) greater than 5 considered to be indicative of problematic multicollinearity.

## RESULTS

### Study population

Of the 34,013 patients, 13,276 patients (median [IQR] age=58.2 [47.20, 66.20], 69.9% male) met the inclusion criteria **(FIGURE I)**. The median [IQR] time on ECMO support was 119 [65, 199] hours. In total, 4.0% (n=525) of patients developed ABI (2.3% (n=307) IS, 1.1% (n=145) ICH, 0.6% (n=73) HIBI). Hospital mortality was 50.5% (n=6,709) **(SUPPLEMENTAL TABLE II)**. 1,456 (11.0%) patients received LV venting while on VA-ECMO. Of these patients, 954 (65.5%) received mAFP, 436 (29.9%) received IABP, and 66 (4.6%) received another type of LV venting. **Figure II** shows the proportions of ABI subtypes stratified by LV venting procedure type. Patients who received LV venting were more likely to be male (73% vs. 69.5%, p=0.006). LV-vented patients also spent longer on ECMO (median [IQR] hours=139.5 [86, 216] vs. 117 [64, 196], p<0.001] and had more RRT (30.8% vs. 25.8%, 0<0.001), hemolysis (7.8% vs. 3.6%, p<0.001) and arrhythmia (20.7% vs. 14.5%, p<0.001) **(TABLE I)**.

**Fig. I.**
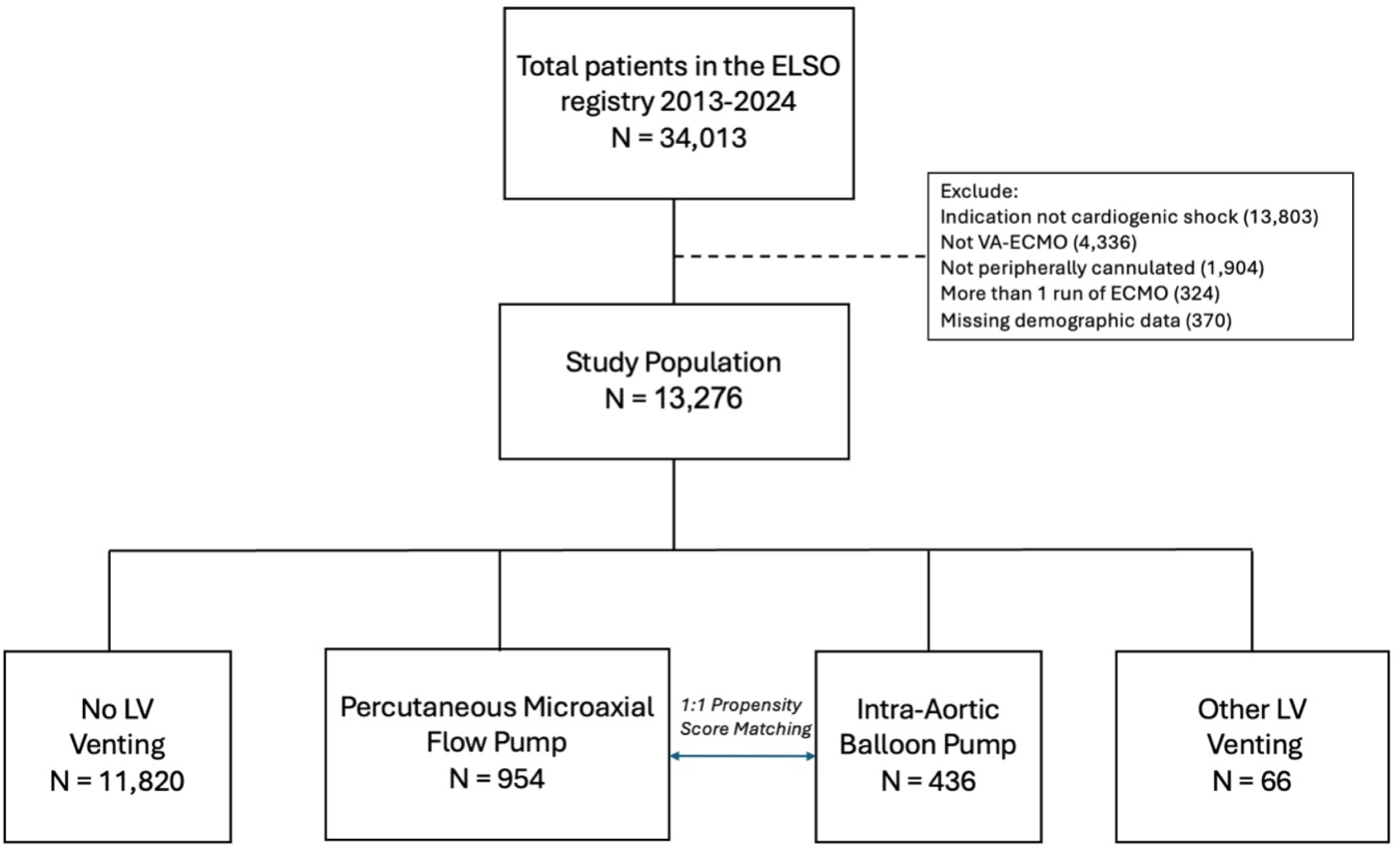
CONSORT Diagram for the Study Cohort; ELSO, Extracorporeal Life Support Organization; VA, venoarterial; ECMO, extracorporeal membrane oxygenation; LV, left ventricular

**Fig. II.**
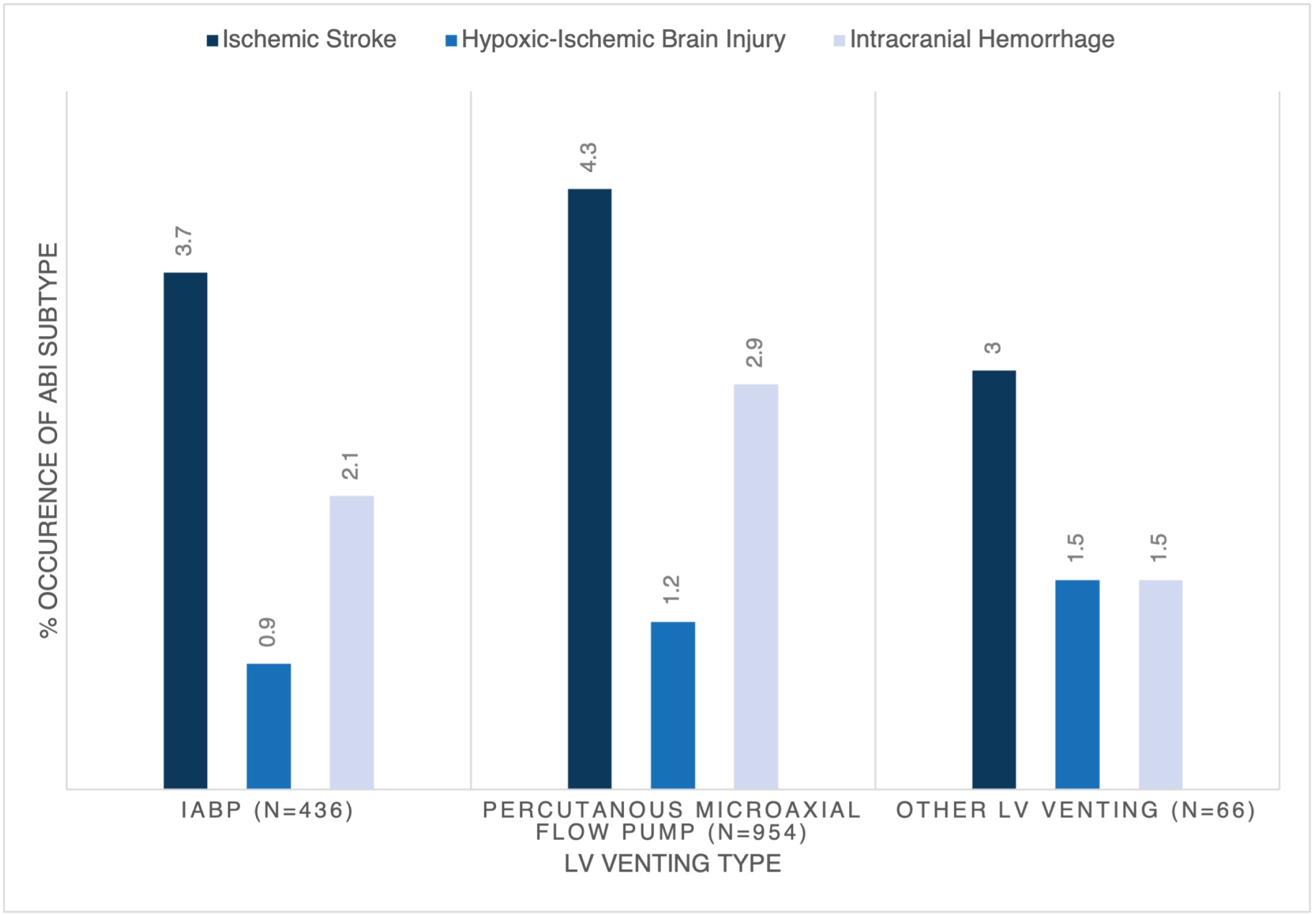
Distribution of ABI for LV Venting Patients Stratified by Procedure Type; IABP, intra-aortic balloon pump; LV, left ventricular

**TABLE I.** Patient Characteristics Stratified by LV Venting.

|  | No LV venting<br>(n=11,820) | LV venting (n=1,456) | p-value |
| --- | --- | --- | --- |
| **Demographics** |  |  |  |
| Age (median [IQR]) | 58.2 [47.2, 66.3] | 57.7 [46.4, 65.6] | 0.177 |
| Male Sex (%) | 8213 (69.5) | 1063 (73.0) | 0.006 |
| BMI (median [IQR]) | 28.24 [24.51, 32.88] | 28.98 [25.31, 33.28] | 0.001 |
| **Hours on ECMO (median [IQR])** | 117 [64.0, 196] | 139.5 [86.0, 216] | <0.001 |
| **Ventilation Type (%)** |  |  | <0.001 |
| Conventional | 7615 (87.3) | 913 (82.9) |  |
| HFO | 13 (0.1) | 0 (0.0) |  |
| No Ventilator | 972 (11.1) | 171 (15.5) |  |
| Other | 116 (1.3) | 16 (1.5) |  |
| Other HFV | 2 (0.0) | 1 (0.1) |  |
| **Transplant (%)** | 782 (6.9) | 111 (7.7) | 0.33 |
| **Ventilation (median [IQR])** |  |  |  |
| PEEP | 8.0 [5.0, 10.0] | 8.0 [5.0, 10.0] | 0.375 |
| Rate | 18.0 [14.0, 23.0] | 18.0 [15.0, 24.0] | 0.057 |
| FiO<sub>2</sub> | 100 [60.0, 100] | 100 [60.0, 100] | 0.714 |
| PaO<sub>2</sub> | 104 [73.0, 191] | 111 [74.0, 204] | 0.122 |
| pH | 7.29 [7.19, 7.38] | 7.30 [7.19, 7.39] | 0.590 |
| Delta PCO<sub>2</sub> | -2.0 [-10.2, 6.0] | -1.0 [-10.0, 7.0] | 0.093 |
| **Outcomes (%)** |  |  |  |
| RRT Required | 3054 (25.8) | 449 (30.8) | <0.001 |
| Hemolysis | 427 (3.6) | 113 (7.8) | <0.001 |
| Arrhythmia | 1714 (14.5) | 302 (20.7) | <0.001 |
| GI Hemorrhage | 565 (4.8) | 85 (5.8) | 0.089 |
| ABI | 412 (3.6) | 113 (7.8) | <0.001 |
| Ischemic Stroke | 248 (2.1) | 59 (4.1) | <0.001 |
| Intracranial Hemorrhage | 107 (0.9) | 38 (2.6) | <0.001 |
| HIBI | 57 (0.5) | 16 (1.1) | 0.005 |
| Mortality | 5980 (50.6) | 729 (50.1) | 0.73 |

LV, left ventricular. BMI, body mass index. ECMO, extracorporeal membrane oxygenation. HFO, high frequency oscillatory. PEEP, positive end-expiratory pressure. FiO<sub>2</sub>, fraction of inspired oxygen. PaO<sub>2</sub>, partial pressure of oxygen in arterial blood. PCO<sub>2</sub>, partial pressure of carbon dioxide in arterial blood. ABI, acute brain injury. HIBI, hypoxic ischemic brain injury. RRT, renal replacement therapy. GI, gastrointestinal.

Similar findings emerged when patients were stratified into no LV venting vs. mAFP. Patients who received mAFP were also more likely to be male and spend longer on ECMO (p<0.05). They also had more RRT, hemolysis, and arrhythmia (p<0.05) **(SUPPLEMENTAL TABLE III)**. Patients who received LV venting using IABP were comparable in sex and BMI to their non-LV-vented counterparts, but spent longer on ECMO (p=0.008) and had more RRT and hemolysis (p<0.05) **(SUPPLEMENTAL TABLE IV)**.

### ABI

#### LV venting vs. no LV venting

Of LV-vented patients, 7.8% (n=113) developed ABI compared to 3.6% (n=412) of non-LV-vented patients (p<0.001) **(TABLE I)**. The distribution of different types of ABI stratified by LV venting procedure type is shown in **Figure I**. Compared to non-LV- vented patients, LV-vented patients had higher prevalence of each type of ABI, including IS (4.1% vs. 2.1%, p<0.001), ICH (2.6% vs. 0.9%, p<0.001), and HIBI (1.1% vs. 0.5%, p=0.005) **(TABLE I)**. After adjusting for covariates in the multivariable regression, patients who received LV venting were found to have higher odds of ABI (aOR=1.76, 95%CI=1.29, 2.37, p<0.001), ICH (aOR=2.13, 95%CI=1.25, 3.52, p=0.004), and HIBI (aOR=2.84, 95%CI=1.18, 6.38, p=0.014) **(TABLE II)**.

**TABLE II.** Multivariable Logistic Regression for ABI and Mortality by LV Venting.

| **Outcome (ref: No LV Venting)** | **Odds Ratio** | **95% CI** | **p-value** |
| --- | --- | --- | --- |
| **LV Venting** |  |  |  |
| ABI | 1.76 | 1.29, 2.37 | <0.001 |
| Ischemic stroke | 1.40 | 0.92, 2.06 | 0.101 |
| Intracranial hemorrhage | 2.13 | 1.25, 3.52 | 0.004 |
| HIBI | 2.84 | 1.18, 6.38 | 0.014 |
| Mortality | 1.08 | 0.91, 1.20 | 0.79 |
| **Percutaneous Microaxial Flow Pump** |  |  |  |
| ABI | 1.62 | 1.11, 2.31 | 0.008 |
| Ischemic stroke | 1.27 | 0.76, 2.03 | 0.33 |
| Intracranial hemorrhage | 1.96 | 1.00, 3.58 | 0.04 |
| HIBI | 2.81 | 0.97, 7.01 | 0.04 |
| Mortality | 1.10 | 0.89, 1.28 | 0.39 |
| **IABP** |  |  |  |
| ABI | 2.20 | 1.34, 3.45 | 0.001 |
| Ischemic stroke | 1.76 | 0.90, 3.14 | 0.08 |
| Intracranial hemorrhage | 2.55 | 1.08, 5.30 | 0.02 |
| HIBI | 3.19 | 0.73, 9.89 | 0.071 |
| Mortality | 1.03 | 0.77, 1.37 | 0.86 |

LV, left ventricular. ABI, acute brain injury. HIBI, hypoxic-ischemic brain injury. IABP, intra-aortic balloon pump.

#### mAFP vs. no LV venting

Patients who received mAFP had more ABI than patients who did not receive LV venting while on VA-ECMO (8.4% (n=80) vs. 3.6% (n=412), p<0.001) **(SUPPLEMENTAL TABLE III)**. mAFP patients had higher prevalence of all ABI types including IS (4.3% vs. 2.1%, p<0.001), ICH (2.9% vs. 0.9%, p<0.001), and HIBI (1.2% vs. 0.5%, p<0.013).

Multivariable regression revealed higher odds of ABI (aOR=1.62, 95%CI=1.11, 2.31, p=0.008, ICH (aOR=1.96, 95%CI=1.00, 3.58, p=0.04), and HIBI (aOR=2.81, 95%CI=0.97, 7.01, p=0.04) for mAFP patients compared to non-LV-vented patients **(TABLE II)**.

#### IABP vs. no LV venting

6.7% (n=29) of IABP patients developed ABI compared to 3.6% (n=412) of non-LV- vented patients (p=0.001**) (SUPPLEMENTAL TABLE IV)**. Specifically, patients who received IABP had higher prevalence of ICH (2.1% vs. 0.9%, p=0.029) but not IS or HIBI. After multivariable regression, odds of ABI in IABP patients was 2.20 times as high (aOR=2.20, 95%CI=1.34, 3.45, p=0.001) compared to non-LV-vented patients **(TABLE II)**. Additionally, patients who received IABP were more likely to develop ICH (aOR=2.55, 95%CI=1.08, 5.30, p=0.02).

#### mAFP vs. IABP

Of patients who received IABP, 6.7% (n=29) developed ABI compared to 8.4% (n=80) of patients who received mAFP (p=0.31) **(TABLE III)**. IABP supported patients had lower frequency of ABI subtypes, including IS (3.7% vs. 4.3%, p=0.69), ICH (2.1% vs. 2.9%, 0=0.45), and HIBI (0.9% vs. 1.2%, p=0.91). No differences in odds of ABI or ABI subtype were found after adjusting for covariates **(SUPPLEMENTAL TABLE V)**.

In the propensity-matched cohort (n=514), 10.0% (n=23) of IABP patients developed ABI compared to 7.8% (n=18) of mAFP patients. With respect to ABI subtypes, 5.2% of IABP patients developed IS, 3.5% developed ICH, and 1.3% developed HIBI. In the mAFP group, 3.9% developed IS, 2.6% developed ICH, and 1.3% developed HIBI. Patient characteristics of the mAFP and IABP propensity-matched cohort are provided in **Table III**. The distribution of propensity scores for patients who received mAFP vs. IABP is visualized in **Supplemental Figure I**.

**TABLE III.**
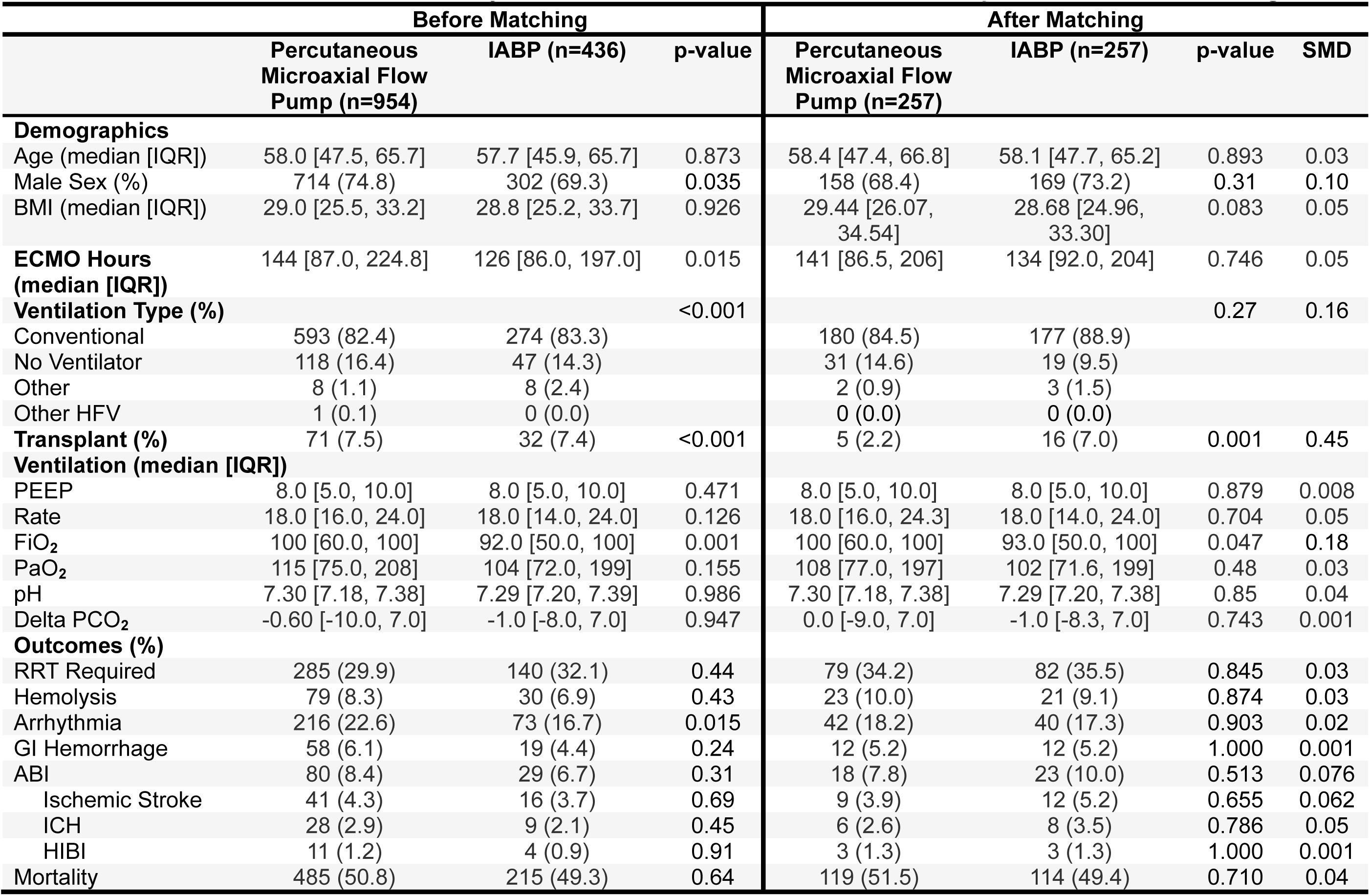
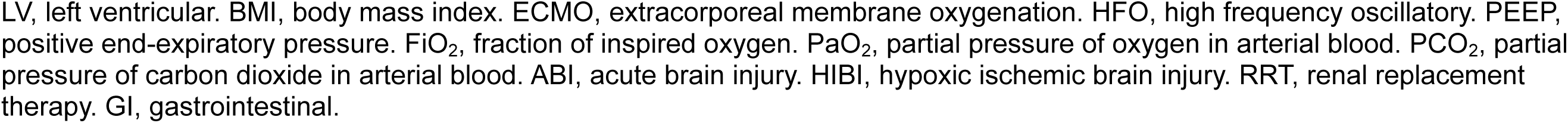
Patient Characteristics Stratified by IABP vs. Percutaneous Microaxial Flow Pump Before and After Matching.

|  | Before Matching |  |  | After Matching |  |  |  |
| --- | --- | --- | --- | --- | --- | --- | --- |
|  | Percutaneous Microaxial Flow Pump (n=954) | IABP (n=436) | p-value | Percutaneous Microaxial Flow Pump (n=257) | IABP (n=257) | p-value | SMD |
| **Demographics** |  |  |  |  |  |  |  |
| Age (median [IQR]) | 58.0 [47.5, 65.7] | 57.7 [45.9, 65.7] | 0.873 | 58.4 [47.4, 66.8] | 58.1 [47.7, 65.2] | 0.893 | 0.03 |
| Male Sex (%) | 714 (74.8) | 302 (69.3) | 0.035 | 158 (68.4) | 169 (73.2) | 0.31 | 0.10 |
| BMI (median [IQR]) | 29.0 [25.5, 33.2] | 28.8 [25.2, 33.7] | 0.926 | 29.44 [26.07, 34.54] | 28.68 [24.96, 33.30] | 0.083 | 0.05 |
| **ECMO Hours (median [IQR])** | 144 [87.0, 224.8] | 126 [86.0, 197.0] | 0.015 | 141 [86.5, 206] | 134 [92.0, 204] | 0.746 | 0.05 |
| **Ventilation Type (%)** |  |  | <0.001 |  |  | 0.27 | 0.16 |
| Conventional | 593 (82.4) | 274 (83.3) |  | 180 (84.5) | 177 (88.9) |  |  |
| No Ventilator | 118 (16.4) | 47 (14.3) |  | 31 (14.6) | 19 (9.5) |  |  |
| Other | 8 (1.1) | 8 (2.4) |  | 2 (0.9) | 3 (1.5) |  |  |
| Other HFV | 1 (0.1) | 0 (0.0) |  | 0 (0.0) | 0 (0.0) |  |  |
| **Transplant (%)** | 71 (7.5) | 32 (7.4) | <0.001 | 5 (2.2) | 16 (7.0) | 0.001 | 0.45 |
| **Ventilation (median [IQR])** |  |  |  |  |  |  |  |
| PEEP | 8.0 [5.0, 10.0] | 8.0 [5.0, 10.0] | 0.471 | 8.0 [5.0, 10.0] | 8.0 [5.0, 10.0] | 0.879 | 0.008 |
| Rate | 18.0 [16.0, 24.0] | 18.0 [14.0, 24.0] | 0.126 | 18.0 [16.0, 24.3] | 18.0 [14.0, 24.0] | 0.704 | 0.05 |
| FiO<sub>2</sub> | 100 [60.0, 100] | 92.0 [50.0, 100] | 0.001 | 100 [60.0, 100] | 93.0 [50.0, 100] | 0.047 | 0.18 |
| PaO<sub>2</sub> | 115 [75.0, 208] | 104 [72.0, 199] | 0.155 | 108 [77.0, 197] | 102 [71.6, 199] | 0.48 | 0.03 |
| pH | 7.30 [7.18, 7.38] | 7.29 [7.20, 7.39] | 0.986 | 7.30 [7.18, 7.38] | 7.29 [7.20, 7.38] | 0.85 | 0.04 |
| Delta PCO<sub>2</sub> | -0.60 [-10.0, 7.0] | -1.0 [-8.0, 7.0] | 0.947 | 0.0 [-9.0, 7.0] | -1.0 [-8.3, 7.0] | 0.743 | 0.001 |
| **Outcomes (%)** |  |  |  |  |  |  |  |
| RRT Required | 285 (29.9) | 140 (32.1) | 0.44 | 79 (34.2) | 82 (35.5) | 0.845 | 0.03 |
| Hemolysis | 79 (8.3) | 30 (6.9) | 0.43 | 23 (10.0) | 21 (9.1) | 0.874 | 0.03 |
| Arrhythmia | 216 (22.6) | 73 (16.7) | 0.015 | 42 (18.2) | 40 (17.3) | 0.903 | 0.02 |
| GI Hemorrhage | 58 (6.1) | 19 (4.4) | 0.24 | 12 (5.2) | 12 (5.2) | 1.000 | 0.001 |
| ABI | 80 (8.4) | 29 (6.7) | 0.31 | 18 (7.8) | 23 (10.0) | 0.513 | 0.076 |
| Ischemic Stroke | 41 (4.3) | 16 (3.7) | 0.69 | 9 (3.9) | 12 (5.2) | 0.655 | 0.062 |
| ICH | 28 (2.9) | 9 (2.1) | 0.45 | 6 (2.6) | 8 (3.5) | 0.786 | 0.05 |
| HIBI | 11 (1.2) | 4 (0.9) | 0.91 | 3 (1.3) | 3 (1.3) | 1.000 | 0.001 |
| Mortality | 485 (50.8) | 215 (49.3) | 0.64 | 119 (51.5) | 114 (49.4) | 0.710 | 0.04 |

LV, left ventricular. BMI, body mass index. ECMO, extracorporeal membrane oxygenation. HFO, high frequency oscillatory. PEEP, positive end-expiratory pressure. $\text{FiO}_2$ , fraction of inspired oxygen. $\text{PaO}_2$ , partial pressure of oxygen in arterial blood. $\text{PCO}_2$ , partial pressure of carbon dioxide in arterial blood. ABI, acute brain injury. HIBI, hypoxic ischemic brain injury. RRT, renal replacement therapy. GI, gastrointestinal.

After propensity matching, odds of ABI remained similar in patients who received IABP and patients who received mAFP (aOR=1.35, 95%CI=0.69, 2.71, p=0.39) **(SUPPLEMENTAL TABLE V)**. Odds of IS (aOR=1.47, 95%CI=0.56, 5.00, p=0.43), ICH (aOR=1.37, 95%CI=0.45, 4.37, p=0.58), and HIBI (aOR=1.24, 95%CI=0.15, 10.7, p=0.83) were also similar between groups.

### Mortality

In-hospital mortality was similar for LV-vented vs non-LV-vented patients (50.1% vs. 50.6%, p=0.73) **(TABLE I)**, mAFP vs. non-LV-vented patients (50.8% vs. 50.6%, p=0.91) **(SUPPLEMENTAL TABLE III),** and IABP vs. non-LV-vented patients (49.3% vs. 50.6%, p=0.63) **(SUPPLEMENTAL TABLE IV)**. After multivariable regression, there were no statistically significant differences in mortality between any LV venting group and the no LV venting group **(TABLE II)**.

In the propensity-matched cohort, in-hospital mortality was 49.4% for patients who received IABP compared to 51.5% for patients who received mAFP (p=0.710).

Odds of mortality for IABP supported patients compared to mAFP supported patients did not differ significantly in the propensity-matched cohort (aOR=0.88, 95%CI=0.58, 1.31, p=0.52) **(SUPPLEMENTAL TABLE V)**.

## DISCUSSION

In this multicenter ELSO registry analysis, we found that LV venting was associated with increased odds of ABI but not mortality in patients receiving peripheral VA-ECMO for cardiogenic shock. Notably, odds of IS were comparable across LV venting, mAFP, and IABP vs. no LV venting groups, while odds of ICH were elevated across each group compared to the no LV venting group. Additionally, odds of HIBI were increased in LV- vented patients compared to non-LV-vented patients. Finally, we found that after propensity matching, there was no significant difference in odds of ABI or mortality for patients who received IABP vs. mAFP during VA-ECMO.

Our results are consistent with the few existing studies that have linked LV venting to increased risk of ABI in patients receiving VA-ECMO, particularly the heightened risk of bleeding complications.^13,16^ Our finding that LV venting was associated with increased odds of ICH but not IS suggests that the primary mechanism of ABI in this clinical context may be related to bleeding risk and hemorrhagic conversion of IS. Notably, LV venting has been found to alter systemic pulsatility; in fact, a prior study in a porcine model showed that ECMO combined with LV support increased carotid artery perfusion compared to ECMO alone, suggesting that changes in pulsatility have the potential to influence cerebral perfusion dynamics.^21^ The absence of pulsatility index measured by transcranial doppler has also been associated with higher rates of intraparenchymal hemorrhage in patients receiving VA-ECMO.^22^ Relatedly, our finding that LV venting was associated with an increased risk of HIBI, despite no significant increase in the risk of IS, may reflect the greater severity of illness in LV-vented patients with the need for LV venting being driven by underlying hemodynamic instability. These patients likely experienced more pronounced hemodynamic fluctuations both before and after venting, contributing to compromised cerebral perfusion and elevated risk of HIBI. The distinctions in risk of particular ABI subtypes underscore the need to better understand the delicate balance between supporting cardiac function and maintaining optimal cerebral perfusion.

Furthermore, our study adds to the literature surrounding LV venting and mortality. One commonly cited study found LV venting to be associated with higher complication rates but lower 30-day mortality in patients with cardiogenic shock.^16^ The international, multicenter, randomized DanGer Shock trial also found improved 180-day mortality in patients receiving mAFP vs. standard of care.^23^ Our study found that despite being associated with increased odds of ABI, hospital mortality was similar between LV- vented and non-LV-vented groups. These findings could represent a delayed mortality benefit associated with LV venting. Given that ABI is typically associated with increased mortality in ECMO patients^24^ and that LV-vented patients tend to be more critically ill, it is possible that LV venting helped mitigate short-term mortality risk.

Taken together, our findings call for a more nuanced approach to patient selection for LV venting. While LV venting may provide valuable hemodynamic support, its potential to increase ABI risk warrants careful consideration, rather than broad, routine application for all patients as suggested by existing literature.^25^. However, our finding that there were no significant differences in odds of ABI or mortality for patients who received IABP vs. mAFP during VA-ECMO reassures clinicians that both mechanical circulatory support devices may offer comparable neurologic safety profiles in this critical patient population. This suggests that treatment decisions can be guided by device availability and patient-specific and physiological factors rather than concerns about differential risks for ABI and mortality. Still, further research is warranted to explore the long-term outcomes for patients supported by either device. Further study on the degree to which the increased ABI risk is driven by LV venting remains necessary.

This study has several limitations. Firstly, the retrospective, observational nature of the dataset limited our ability to infer causality from our findings despite propensity matching and adjustment for known confounders. Secondly, the voluntary nature of the ELSO dataset could have resulted in selection bias, and variations in data reporting between centers may affect generalizability. Missing data from the registry could also have limited our analyses, though we attempted to mitigate this by using listwise deletion of cases with missing covariates for our propensity matching analysis. Thirdly, our study lacked detailed anticoagulation parameters, and we were limited in our ability to control for illness severity. Fourthly, outcomes were limited to hospital stay. Finally, since the registry did not include data on the duration of LV venting support, we were unable to identify patients who had LV venting devices placed pre-ECLS but maintained throughout ECMO support. This gap also limited our ability to evaluate the impact of LV venting duration with respect to our primary and secondary outcomes.

## CONCLUSIONS

LV venting was associated with increased odds of ABI but not mortality in patients receiving peripheral VA-ECMO for cardiogenic shock. Similar mortality between LV- vented and non-LV-vented patients despite increased ABI risk with LV venting may suggest an unmeasured survival benefit of LV venting. There was no difference in odds of ABI or mortality in patients who received IABP vs. mAFP. Further research is essential to validate these findings and better understand the mechanisms linking LV venting, ABI, and survival.

## Data Availability

The data used in this study were obtained from the Extracorporeal Life Support Organization (ELSO) registry, a comprehensive international database of patients receiving extracorporeal life support. Access to the ELSO registry data is available upon request and subject to approval by the ELSO Scientific Oversight Committee. For more information on accessing the registry data, including submission guidelines and requirements, please visit the ELSO website at www.elso.org.

## Non-standard Abbreviations and Acronyms

ABI: acute brain injury.
GI: gastrointestinal.
HFO: high frequency oscillatory.
HIBI: hypoxic ischemic brain injury.
IABP: intra-aortic balloon pump.
IS: ischemic stroke.
LV: left ventricular.
mAFP: microaxial flow pump.
RRT: renal replacement therapy.

